# Immunogenicity and safety of Quadrivalent Influenza HA vaccine compared with Trivalent Influenza HA vaccine and evaluation of Quadrivalent Influenza HA vaccine batch-to-batch consistency in Indonesian children and adults

**DOI:** 10.1101/2023.01.27.23285092

**Authors:** Eddy Fadlyana, Meita Dhamayanti, Rodman Tarigan, Susantina Prodjosoewojo, Andri Reza Rahmadi, Rini Mulia Sari, Kusnandi Rusmil, Cissy B Kartasasmita

**Affiliations:** Department of Child Health, Faculty of Medicine Universitas Padjadjaran/Hasan Sadikin Hospital, Bandung, Indonesia; Department of Internal Medicine, Faculty of Medicine Universitas Padjadjaran/Hasan Sadikin Hospital, Bandung, Indonesia; Surveillance and Clinical Trial Division, PT Bio Farma, Bandung, Indonesia

**Keywords:** children, adult, immunogenicity, Quadrivalent Influenza HA vaccine, safety, Trivalent Influenza HA vaccine

## Abstract

**Background:** One of the newest strategies developed by the Global Influenza Strategy has been to broaden the composition of the current influenza vaccine formulations from trivalent products to quadrivalent products. This study aimed to assess the immunogenicity and safety of Quadrivalent Influenza HA vaccine (QIV) compared with Trivalent Influenza HA vaccine (TIV) and to evaluate three consecutive batches of QIV equivalence in Indonesian children and adults.

**Methods and findings:** This was an open-labeled, bridging clinical study involving unprimed healthy children and adults aged 9–40 years. A total of 540 subjects were enrolled in this study and randomized into four arm groups. Each subject received one dose of TIV or QIV with three different batch codes. Serology tests were performed at baseline and 28 days after vaccination. Hemagglutination inhibition (HI) antibody titers were analyzed for Geometric Mean Titer (GMT), seroprotection, and seroconversion rates. Solicited, unsolicited, and serious adverse events were observed up to 28 days after vaccination. A total of 537 subjects completed the study per protocol and were analyzed for immunogenicity criteria. All randomized subjects were analyzed for safety criteria. The percentage of the subjects with anti-HI titer ≥1:40 28 days after QIV vaccination was 99.5% for A/H1N1; 99.5% for A/H3N2; 93.1% for B/Texas, and 99.0% for B/Phuket. The seroprotection, GMT, and seroconversion rates of QIV were not significantly different from those of TIV for the common vaccine strains (*p* > 0.01) and were significantly different from those of TIV for the added B/Phuket strains (*p* < 0.01). Most solicited injection-site and systemic reactions with either vaccine were mild to moderate and resolved within a few days.

**Conclusions:** QIV was immunogenic and well-tolerated and had immunogenicity and safety profiles compared with TIV for all common strains. The immunogenicity of the three batches of QIV was equivalent for the four strains.

**Clinical Trial registration:** NCT03336593

## Introduction

Influenza is an acute viral illness of the respiratory tract and poses a substantial public health burden in terms of morbidity, mortality, and costs. The World Health Organization (WHO) reported that 3–5 million cases of severe influenza occur each year worldwide, resulting in about 290,000 to 650,000 related deaths per year. One of the goals of the Global Influenza Strategy for 2019–2030 is to reduce the burden of seasonal influenza by promoting research and innovation for improved influenza vaccines [1]. Annual influenza immunization is recommended in elderly subjects, children aged six months or more, pregnant women, and individuals with chronic conditions, such as respiratory/heart/liver diseases, diabetes, or a weakened immune system. These categories are at heightened risk of influenza-related complications and mortality [2].

Internationally available vaccines for controlling seasonal influenza are safe and effective and have the potential to prevent significant annual morbidity and mortality. The WHO annually recommends composing vaccines based on global virological surveillance. Annual Trivalent influenza HA vaccines (TIVs) contain two influenza A strains (H1N1 and H3N2) and only one influenza B virus [3]. Therefore, the effectiveness of TIVs depends on the degree of matching between the vaccine strain and circulating viral strains. In the last two decades, four major and at least eight minor mismatches between vaccine and circulating B viruses have occurred in the northern hemisphere, thus impairing the performance of TIVs [4]. Specifically, Ambrose et al. observed that a B-mismatch between vaccine and circulating strains occurred in Europe in 5 of 10 seasons between 2001 and 2011 [5].

In February 2009, the Food and Drug Administration (FDA), for the first time, considered the inclusion of an additional influenza B strain in the antigenic composition of seasonal influenza vaccines to minimize the impact of B strain-mismatch on vaccine effectiveness [6]. Subsequently, in February 2012, the WHO recommended the production of Quadrivalent Influenza HA vaccines (QIVs) for seasonal immunization. In 2012, the European Medicines Agency (EMA) also highlighted the need for a quadrivalent vaccine that could overcome the lack of protection against the influenza B lineage not present in the trivalent vaccine. Finally, in February 2013, the WHO issued its first guidelines recommending that both expected B strains be included in the vaccine composition [7,8].

Bio Farma conducted several studies on seasonal TIVs between 2008 and 2014. The results of these studies were consistent. TIVs were well-tolerated and induced high antibody titer against influenza antigens and no serious adverse events (AEs) during the study [9–13]. This was the first QIV study conducted by Bio Farma on subjects aged 9–40 years in Indonesia. QIVs that could potentially provide wider protection against influenza B viruses are becoming available, and recommendations should not be limited to trivalent vaccine formulations [3]. This study aimed to assess the immunogenicity and safety of QIV compared with TIV and to evaluate batch-to-batch consistency in three consecutive batches of QIV.

## Materials and methods

### 2.1 Study design

This was an experimental open-labeled, four arm bridging study. The study was a collaboration between the Department of Child Health, Faculty of Medicine, Universitas Padjadjaran, and PT Bio Farma (Persero), Indonesia. This study was approved by the Research Ethics Committee of the Faculty of Medicine, Universitas Padjadjaran, and conducted in accordance with the Declaration of Helsinki and the International Conference on Harmonization Good Clinical Practice guidelines. Written informed consent was obtained from the participants or their parents before performing any study-specific procedure.

### 2.2 Study subjects

A total of 540 subjects were enrolled in this study. Subjects were enrolled from 3 primary care centers in Bandung City, Ibrahim Adjie Primary Health Center, Puter Primary Health Center, and Garuda Primary Health Center, from October 2017 to June 2018. The primary inclusion criteria included healthy children and adults aged 9–40 years committed to complying with study instructions and trial schedules. Subjects were not eligible if they presented with mild, moderate, or severe illness with a fever (axillary temperature ≥37.5°C). Other exclusion criteria included allergic to egg, chicken protein, or other vaccine components and a history of blood disorders contraindicating intramuscular injection.

### 2.3 Randomization and blinding

For each subject recruited, the inclusion number was allocated in the chronological order of the subject, which was included in the trial from I-001 to I-180 (for the 9–12-year age group), II-001 to II-180 (for the 13–17-year age group), and III-001 to III-180 (for the 18–40-year age group). The subjects were randomized into treatment groups. The doctor strictly followed the list of randomization provided by Bio Farma. Treatment was allocated in accordance with a randomization list so that each randomization number corresponded to only one strictly randomly assigned treatment group (QIV batch A, QIV batch B, QIV batch C, and TIV).

### 2.4 Vaccines and vaccination schedule

The QIV vaccine was formulated by PT Bio Farma (Persero), Indonesia, using bulks imported from Japan. The investigational QIV contained 15 *μ* HA from each of 4 strains, A/California/7/2009 (X-179A) (H1N1) pdm09n, A/Hong Kong/4801/2014 (X-263) (H3N2), B/Texas/2/2013, and B/Phuket/3073/2013, in a 0.5 ml dose, with batch numbers A: 3070117, B: 3070217, and C: 3070317. TIV contained 15 *μ*g HA of each of 3 strains, A/California/7/2009 (X-179A) (H1N1) pdm09n, A/Hong Kong/4801/2014 (X-263) (H3N2), and B/Texas/2/2013, in 0.5 ml dose. Each subject received one dose (0.5 ml) of TIV or QIV with different batch numbers: 3070117, 3070217, and 3070317 for batches A, B, and C, respectively, according to the randomization.

#### Sample size and study analysis

The sample size was determined based on a 95% confidence interval (CI) and a test power of 80%. The required sample size was 115 in each group, with 10% dropout anticipation. With the assumption that not all the subjects could complete the study, the total number of subjects was added at least 10% from the minimum requirement (1/1 − 0.1) × 115 = 128. Demographic data were expressed as mean, standard deviation, and range values. Immunogenicity analyses were performed on the per protocol population. Analysis of Geometric Mean Titer (GMT), seroprotection, and seroconversion rates between the vaccine groups was performed using the Chi-square, McNemar, or Wilcoxon tests. Values of *p* < 0.05 indicated statistically significant differences between groups.

The safety analyses were based on the intention-to-treat population analyses. The safety data were collected up to 28 days after the vaccination. The subjects were provided with a diary card to record the appearance, duration, and intensity (mild, moderate, or severe) of any solicited AE (local pain, redness, swelling, induration, fever, fatigue, and myalgia) and unsolicited AE. Local pain was graded as mild (mild pain at the injection-site when touched), moderate (pain with movements), and severe (significant pain at rest). Redness, induration, and swelling intensity were measured using a plastic bangle and categorized as mild (<5 cm), moderate (5–10 cm), and severe (>10 cm). Fever was graded as mild (38.0°C–38.4°C), moderate (38.5°C– 38.9°C), and severe (≥39.0°C). Fatigue, myalgia, and unsolicited events were graded as mild (no interference with activity), moderate (some interference with activity), and severe (prevents daily activity, requires medical intervention).

## Results

### 3.1 Demographics and baseline characteristics

In this study, 405 and 135 subjects were enrolled in the QIV and TIV groups, respectively. Of the 405 subjects in the QIV group, three were excluded from immunogenicity analysis because of protocol non-compliance (Fig 1). The demographic characteristics of study participants showed a fair distribution in gender and age (Table 1).

**Fig 1.**
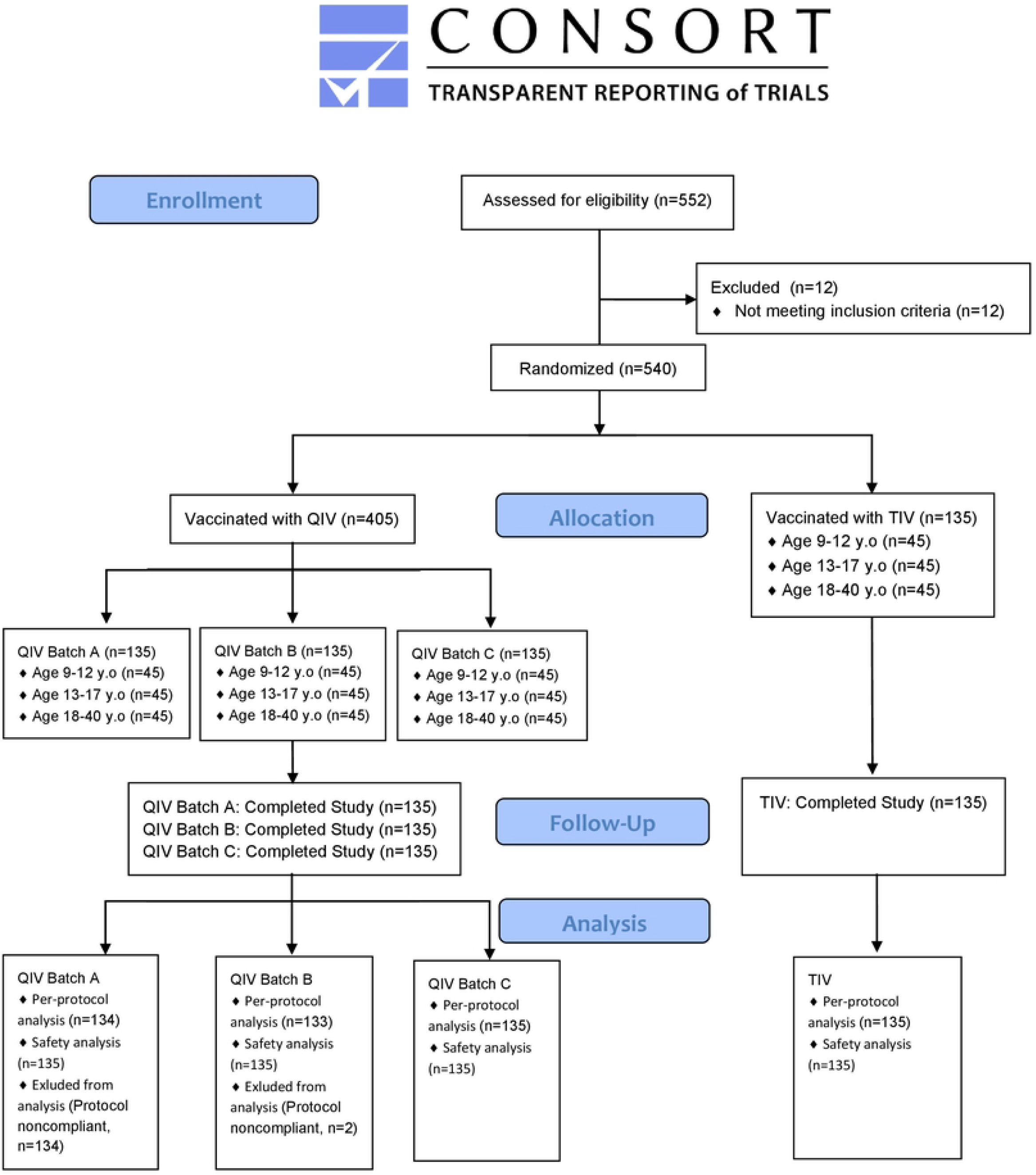
Participant disposition.

**Table 1.** Demographic characteristics of study participants.

| Charateristics | Batch A<br>(n = 135) | Batch B<br>(n = 135) | Batch C<br>(n = 135) | TIV<br>(n = 135) |
| --- | --- | --- | --- | --- |
| Sex, n (%) |  |  |  |  |
| Male | 67 (49.6) | 67 (49.6) | 57( 42.2) | 64 (47.4) |
| Female | 68 (50.4) | 68 (50.4) | 78 (57.8) | 71 (52.6) |
| Age (y) |  |  |  |  |
| Mean ± standard deviation | 17.21 ± 7.64 | 17.70 ± 8.48 | 17.74 ± 8.31 | 17.18 ± 8.27 |
| Median | 15 | 14 | 14 | 14 |
| Range | 9–39 | 9–40 | 9–40 | 9–39 |

### 3.2 Immunogenicity

Table 2 shows a comparison of the percentage of subjects with anti-HI titer ≥1:40 28 days after QIV and TIV. No difference in seroprotection was observed between one dose of QIV and TIV, except for B/Phuket/3073/2013.

**Table 2.**
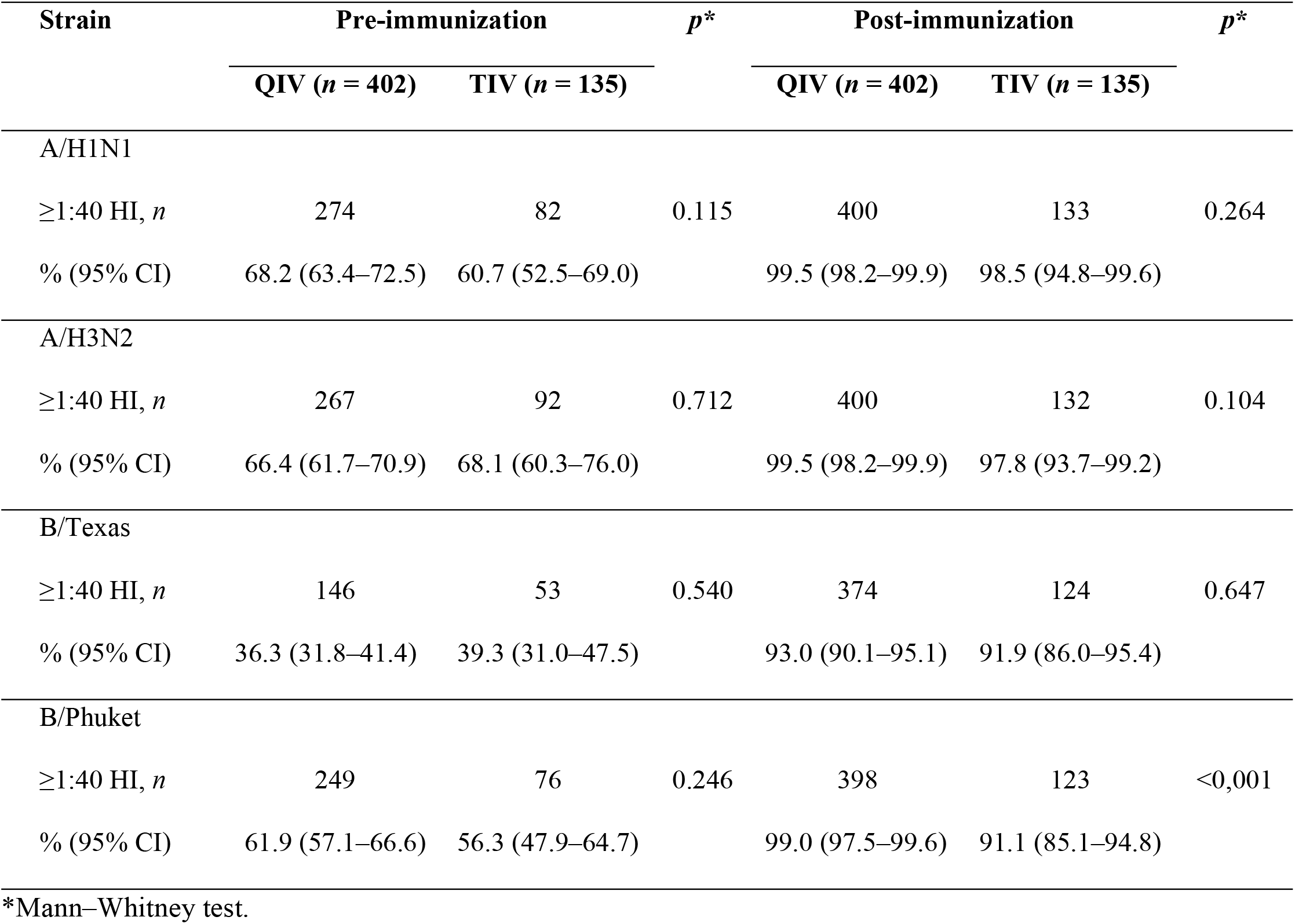
Influenza seroprotection rate before and 28 days after immunization.

| Strain | Pre-immunization |  | *p* * | Post-immunization |  | *p* * |
| --- | --- | --- | --- | --- | --- | --- |
|  | QIV (*n* = 402) | TIV (*n* = 135) |  | QIV (*n* = 402) | TIV (*n* = 135) |  |
| A/H1N1 |  |  |  |  |  |  |
| ≥1:40 HI, *n* | 274 | 82 | 0.115 | 400 | 133 | 0.264 |
| % (95% CI) | 68.2 (63.4–72.5) | 60.7 (52.5–69.0) |  | 99.5 (98.2–99.9) | 98.5 (94.8–99.6) |  |
| A/H3N2 |  |  |  |  |  |  |
| ≥1:40 HI, *n* | 267 | 92 | 0.712 | 400 | 132 | 0.104 |
| % (95% CI) | 66.4 (61.7–70.9) | 68.1 (60.3–76.0) |  | 99.5 (98.2–99.9) | 97.8 (93.7–99.2) |  |
| B/Texas |  |  |  |  |  |  |
| ≥1:40 HI, *n* | 146 | 53 | 0.540 | 374 | 124 | 0.647 |
| % (95% CI) | 36.3 (31.8–41.4) | 39.3 (31.0–47.5) |  | 93.0 (90.1–95.1) | 91.9 (86.0–95.4) |  |
| B/Phuket |  |  |  |  |  |  |
| ≥1:40 HI, *n* | 249 | 76 | 0.246 | 398 | 123 | <0,001 |
| % (95% CI) | 61.9 (57.1–66.6) | 56.3 (47.9–64.7) |  | 99.0 (97.5–99.6) | 91.1 (85.1–94.8) |  |

\*Mann–Whitney test.

Fig 2 shows a comparison of GMT between subjects who received QIV and TIV. No significant differences in GMT were observed between one dose of QIV and TIV in the 9–40-year-old subjects for A/California/7/2009 (X-179A) (H1N1) pdm09, A/Hong Kong/4801/2014 (X-263) (H3N2), and B/Texas/2/2013 (*p* = 0.322, *p* = 0.536, and *p* = 0.378, respectively). However, a significant difference in GMT was observed between one dose of QIV and TIV for strain B/Phuket/3073/2013 (*p* < 0.001).

**Fig 2.**
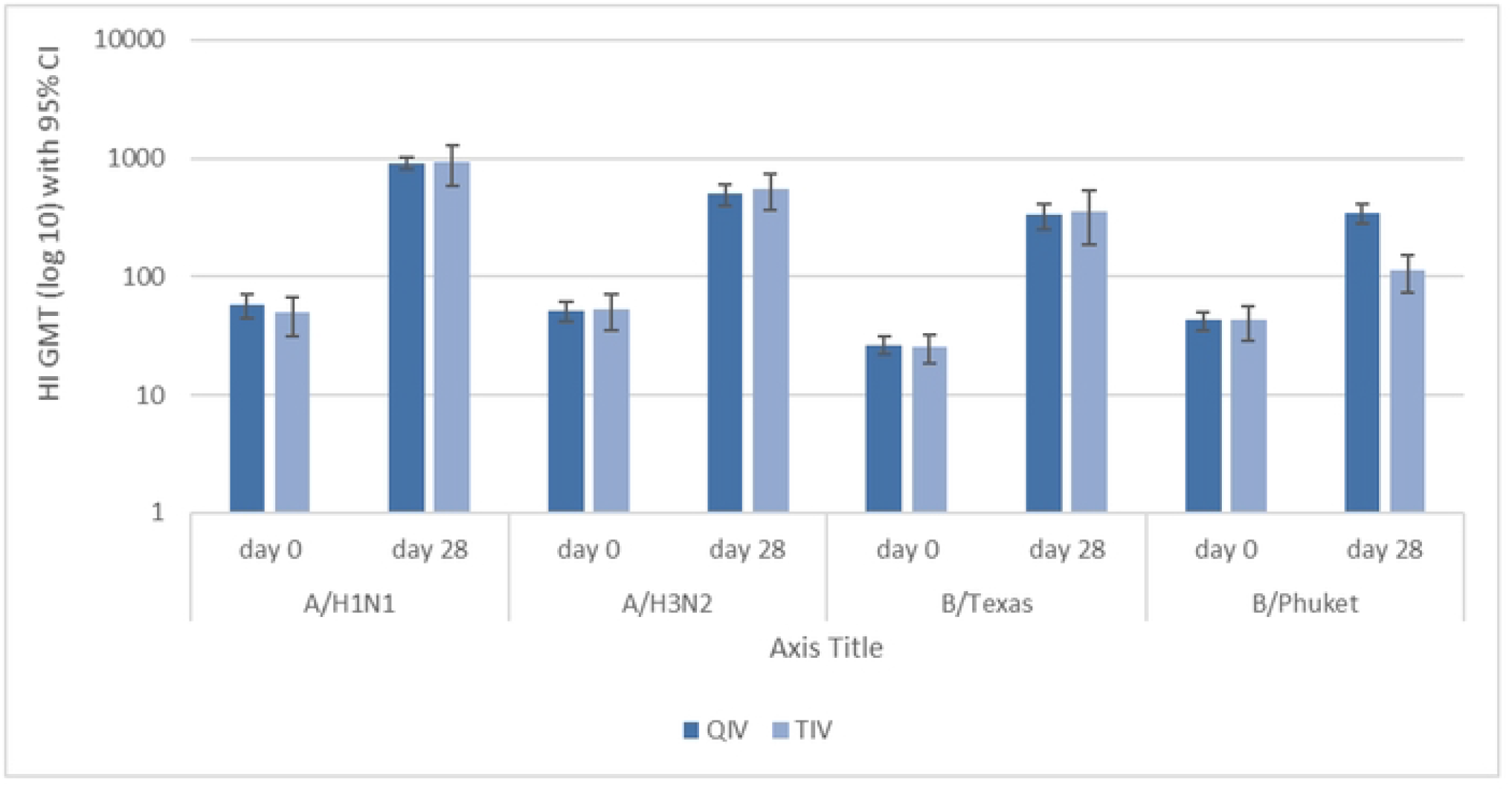
Hemagglutination inhibition (HI) Geometric Mean Titers (GMT) before and 28 days after QIV and TIV immunization.

The transition from seronegative to seropositive is defined as a pre-vaccination titer <1:40 HI units and a post-vaccination titer ≥1:40 HI units. Seroconversion was also defined as increasing antibody titer ≥4 times and transition from seronegative to seropositive (Table 3).

**Table 3.** Differences in seroconversion rates.

| Strain | Increasing antibody titer |  |  |  | Transition from seronegative to seropositive<sup>b</sup> |  |  |  |
| --- | --- | --- | --- | --- | --- | --- | --- | --- |
| | ≥4 times<sup>a</sup> | | | | | | | |
|  | *n* (%) |  |  |  | Diff (%)<sup>c</sup> |  |  |  |
|  | QIV | TIV | %Diff | *p* | QIV | TIV | Diff | *p*<sup>d</sup> |
|  |  |  | (95% CI) |  |  |  | (95% CI) |  |
| A/H1N1 | 333 (82.2) | 115 (85.2) | -3.0 (-9.4; 4.8) | 0.525 | 98.4% | 96.2% | 2.2 (-0.8; 6.7) | 0.582 |
| A/H3N2 | 331 (81.7) | 113 (83.1) | -1.4 (-8.1; 6.6) | 0.717 | 98.5% | 93.0% | 5.5 (1.6; 10.8) | 0.092 |
| B/Texas | 349 (86.1) | 114 (84.4) | 1.7 (-4.6; 9.4) | 0.489 | 89.1% | 86.6% | 2.5 (-3.4; 9.7) | 0.541 |
| B/Phuket | 335 (82.7) | 56 (41.5) | 41.2 (31.9; 49.9) | <0.001 | 97.4% | 79.7% | 17.7 (11.0; 24.9) | <0.001 |

CI, confidence interval.
<sup>a</sup> Number of subjects (*n*) on increasing antibody titer for group QIV = 402 and group TIV = 135.
<sup>b</sup> Number of subjects (*n*) on transition from seronegative to seropositive for each group, and each strain was based on the number of seronegative subjects at baseline (pre-vaccination).
<sup>c</sup> Percentage (%) defined as the percentage of subjects with anti-HI titer <1:40 HI (seronegative) at baseline and ≥ 1:40 HI (seropositive) after vaccination.
<sup>d</sup> *p* value based on the Fisher exact test

Batch-to-batch equivalence for each strain was concluded if the two-sided 95% CI of each strain GMT ratio of the compared batches was between 0.67 and 1.5 (Table 4) [14].

**Table 4.** QIV batch-to-batch comparison.

| Age group | Strain | Comparison | GMT | Equivalence |
| --- | --- | --- | --- | --- |
|  |  |  | post-vaccination |  |
|  |  |  | ratio (95% CI) |  |
| 9–12 years | A/H1N1 | Batch A vs. batch B | 1.030 (0.996–1.061) | Yes |
| *n* = 135 |  | Batch A vs. batch C | 1.020 (0.976–1.072) | Yes |
|  |  | Batch B vs. batch C | 1.082 (0.988–1.176) | Yes |
|  | A/H3N2 | Batch A vs. batch B | 1.074 (0.996–1.152) | Yes |
|  |  | Batch A vs. batch C | 1.027 (0.971–1.083) | Yes |
|  |  | Batch B vs. batch C | 0.987 (0.928–1.045) | Yes |
|  | B/Texas | Batch A vs. batch B | 0.951(0.941–1.061) | Yes |
|  |  | Batch A vs. batch C | 1.003 (0.867–1.138) | Yes |
|  |  | Batch B vs. batch C | 1.107 (0.979–1.236) | Yes |
|  | B/Phuket | Batch A vs. batch B | 1.076 (1.004–1.147) | Yes |
|  |  | Batch A vs. batch C | 1.089 (1.019–1.159) | Yes |
|  |  | Batch B vs. batch C | 1.045 (0.960–1.129) | Yes |
| 13–17 years<br>*n* = 132 | A/H1N1 | Batch A vs. batch B | 1.010 (0.958–1.063) | Yes |
|  |  | Batch A vs. batch C | 0.981 (0.938–1.023) | Yes |
|  |  | Batch B vs. batch C | 0.982 (0.944–1.021) | Yes |
|  | A/H3N2 | Batch A vs. batch B | 0.997 (0.925–1.068) | Yes |
|  |  | Batch A vs. batch C | 1.023 (0.949–1.096) | Yes |
|  |  | Batch B vs. batch C | 1.053 (0.984–1.121) | Yes |
|  | B/Texas | Batch A vs. batch B | 1.025 (0.916–1.133) | Yes |
|  |  | Batch A vs. batch C | 0.972 (0.881–1.064) | Yes |
|  |  | Batch B vs. batch C | 1.000 (0.913–1.087) | Yes |
|  | B/Phuket | Batch A vs. batch B | 1.009 (0.948–1.070) | Yes |
|  |  | Batch A vs. batch C | 0.971 (0.909–1.034) | Yes |
|  |  | Batch B vs. batch C | 0.984 (0.913–1.055) | Yes |
| 18–40 years<br>*n* = 135 | A/H1N1 | Batch A vs. batch B | 1.051 (0.982–1.120) | Yes |
|  |  | Batch A vs. batch C | 1.073 (0.996–1.150) | Yes |
|  |  | Batch B vs. batch C | 1.036 (0.975–1.097) | Yes |
|  | A/H3N2 | Batch A vs. batch B | 1.123 (1.034–1.211) | Yes |
|  |  | Batch A vs. batch C | 1.062 (0.968–1.155) | Yes |
|  |  | Batch B vs. batch C | 0.985 (0.889–1.081) | Yes |
|  | B/Texas | Batch A vs. batch B | 1.028 (0.927–1.129) | Yes |
|  |  | Batch A vs. batch C | 1.064 (0.958–1.169) | Yes |
|  |  | Batch B vs. batch C | 1.067 (0.969–1.165) | Yes |
|  | B/Phuket | Batch A vs. batch B | 1.065 (0.987–1.144) | Yes |
|  |  | Batch A vs. batch C | 1.084 (0.998–1.170) | Yes |
|  |  | Batch B vs. batch C | 1.042 (0.967–1.117) | Yes |
| All age | A/H1N1 | Batch A vs. batch B | 1.029 (0.996–1.061) | Yes |
| (9–40 years) |  | Batch A vs. batch C | 1.046 (1.003–1.089) | Yes |
| *n* = 402 |  | Batch B vs. batch C | 1.030 (0.989–1.070) | Yes |
|  | A/H3N2 | Batch A vs. batch B | 1.066 (1.020–1.111) | Yes |
|  |  | Batch A vs. batch C | 1.037 (0.994–1.080) | Yes |
|  |  | Batch B vs. batch C | 1.008 (0.964–1.051) | Yes |
|  | B/Texas | Batch A vs. batch B | 1.001 (0.941–1.061) | Yes |
|  |  | Batch A vs. batch C | 1.014 (0.950–1.078) | Yes |
|  |  | Batch B vs. batch C | 1.059 (0.998–1.119) | Yes |
|  | B/Phuket | Batch A vs. batch B | 1.050 (1.010–1.090) | Yes |
|  |  | Batch A vs. batch C | 1.049 (1.006–1.091) | Yes |
|  |  | Batch B vs. batch C | 1.024 (0.980–1.067) | Yes |

Age 9–12 years: *n* = 45 for each QIV batch A, B, and C group.
Age 13–17 years: *n* = 44 for QIV batch A group, *n* = 43 for QIV batch B group, and *n* = 45 for QIV batch C group.
Age 18–40 years: *n* = 45 for each QIV batch A, B, and C group.

### 3.3 Safety

Solicited and unsolicited post-vaccination AEs were categorized as immediate (within 30 min), intermediate (30 min to 72 h), and delayed (72 h to 28 days) reactions (Figs 3 and 4).

**Fig 3.**
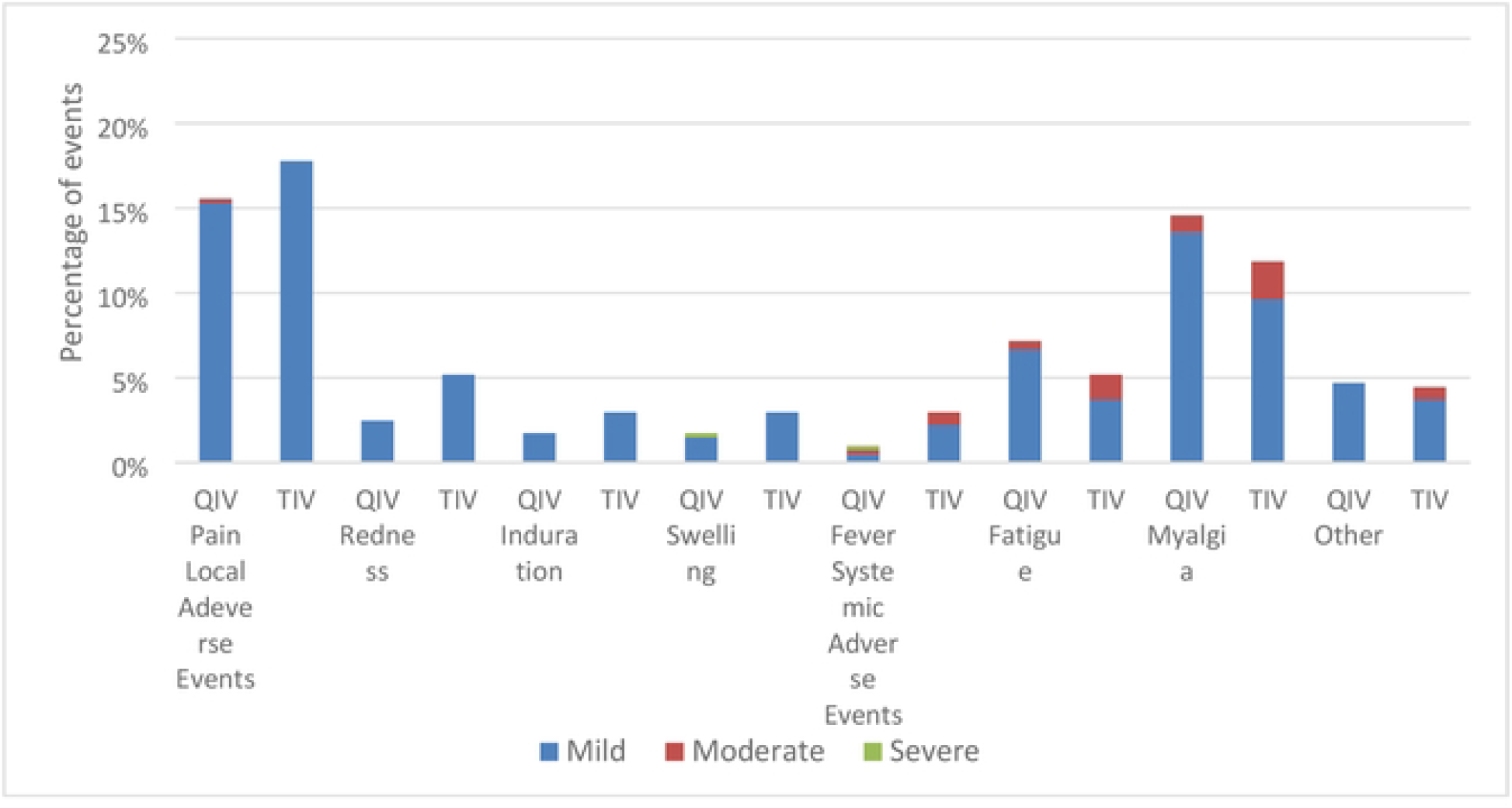
Intensity of reported local and systemic adverse events (AEs).

**Fig 4.**
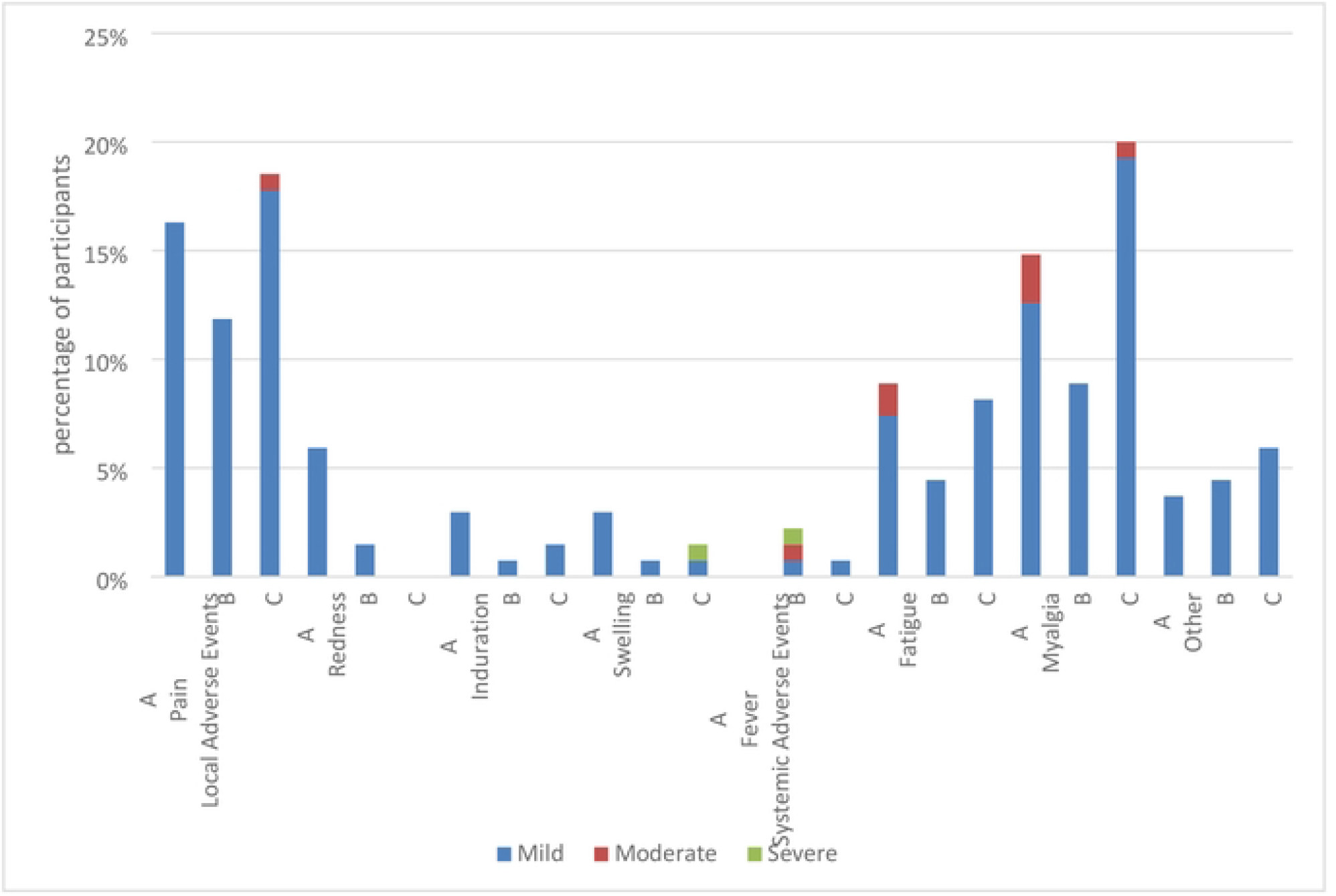
Intensity of reported local and systemic AEs on each QIV batch.

Most local and systemic AEs reported in the QIV and TIV groups had mild intensity. Mild pain was the most local adverse reaction, which occurred in 15.3% and 17.8% of the subjects in the QIV and TIV groups, respectively. Mild myalgia was the most systemic adverse reaction, which occurred in 13.6% and 9.6% of the subjects in the QIV and TIV groups, respectively.

## Discussion

This study was conducted on subjects aged 9–40 years, while a previous study by Dhamayanti et al. was conducted on infants to children aged 8 years [15]. The HI data showed a strong serological response for each of the shared influenza strains in the QIV and TIV groups and the percentage of subjects in the QIV group achieving a serum HI titer ≥40. In this study, in the age group of 9–40 years, the QIV induced comparable immune responses to TIV for A strains and the B lineage common to both QIV and TIV. The protectivity/seroprotection rate of QIV was defined as the percentage of participants with an HI titer ≥40 for A/California/7/2009 (X-179A) (H1N1) pdm09, A/Hong Kong/4801/2014 (X-263) (H3N2), B/Texas/2/2013, and B/Phuket/3073/2013 of QIV (99.5%; 99.5%; 93.1%; 99.0%) and TIV (98.5%; 97.8%; 91.9%; 91.1%). The immunogenicity based on seroprotection rates of a candidate QIV was not significantly different from TIV for shared vaccine strains (*p* > 0.01) and was significantly different from TIV with respect to the added B strains (B/Phuket/3073/2013) (*p* < 0.01). The same result was observed for the GMT as well.

The inclusion of a fourth strain in QIV did not interfere with the EMA criteria for immune responses in adult vaccine recipients. In adults, post-vaccination seroprotection rates were ≥99%, seroconversion rates were >59%, and post-vaccination/pre-vaccination GMT ratios were ≥7.3 for all four vaccine strains. These data demonstrate that the presence of a second influenza B strain in QIV does not negatively affect the immune response to the other strains. Moreover, the immune responses to all strains contained in the two vaccines were robust, with the highest responses to the A/California/7/2009 (H1N1) strain.

The seroconversion rates regarding the percentage of subjects with increasing antibody titer ≥4 times and transition of seronegative to seropositive and of QIV were not significantly different from those of TIV. The superior QIV immunogenicity is expected to correspond with superior protection against influenza B relative to TIV in a season when there is lineage mismatch or cocirculation of two influenza B lineages. These findings are similar to those of the meta-analysis study by Moa in 2015, which confirmed that the inactivated QIV used in the studies in adults had similar efficacy against the three strains shared in common with the TIV (A/H1N1, A/H3N2, and the B lineage included in the TIV) and statistically significant superior efficacy against the B lineage not included in the TIV. The presence of a second influenza B strain in QIV did not negatively affect the immune response to the other strains. The addition of a second B strain in QIV might enhance the protective efficacy of influenza vaccines as it would reduce the undesirable mismatch between the recommended B strain for TIV and the one predominantly circulating [16–17].

This study also showed that the immunogenicity of the three batches of QIV was equivalent for the four strains. Batch-to-batch equivalence of all three batches of QIV was demonstrated for all four strains. The seroprotection rates of three batches for A/California/7/2009 (X-179A) (H1N1) pdm09, A/Hong Kong/4801/2014 (X-263) (H3N2), B/Texas/2/2013, and B/Phuket/3073/2013 were (99.3; 100; 99.3), (99.3; 99.2; 100), (88.8; 95.5; 94.8), and (98.5; 100; 98.5), respectively. The increase in the overall post-vaccination GMTs for each pair of batches for each strain was not different.

During this study, no serious AEs related to the vaccine were observed. This study found comparable reactogenicity and safety profiles between the QIV candidate and the TIV in both adult and adolescent groups. Both vaccines were well-tolerated by both age groups. Local and systemic reactogenicity profiles were also similar between the vaccine groups. Most reactions were mild or moderate in severity and lasted for 1–3 days. Most solicited injection-site and systemic reactions with either vaccine were mild to moderate and resolved within a few days.

Injection-site pain was the most frequently reported solicited local event, and fatigue and myalgia were the commonly reported solicited systemic events among the studies. A meta-analysis study by Moa in 2015 showed a comparable safety profile between QIV and TIV. However, QIV had a slightly increased frequency of injection-site pain compared with TIV, and no statistically significant difference was observed in the overall rate of AEs [16].

The QIV and TIV groups showed similar rates of systemic AEs. In both vaccine groups, mild myalgia was the most frequent systemic AE. These findings are similar to those reported by Wang et al. [18]. The incidence of fever was similar with both vaccines, which is consistent with the results of a previous study [19].

Frequencies of unsolicited AEs in the 28 days following vaccination with QIV were similar between the adult and adolescent groups. These data are consistent with the results of a meta-analysis of five randomized clinical trials, demonstrating no significant difference between QIV and TIV in terms of the frequency of aggregated local and systemic AEs within 7 days after vaccination [16]. The trials reported that the rate of both local and systemic AEs was transient, short-lived, and resolved within 1–3 days in both vaccine groups. No vaccine-related serious AEs or deaths were associated with the vaccines. Although QIV had a slight increase in local reaction (injection-site pain) compared with TIV, the potential benefit of QIV is considered greater with regard to improved protection from infection in the population [16].

QIV and TIV have similar reactogenicity and AE profiles, with no apparent adverse effects on the tolerability of the higher antigen content in QIV (60 μg HA for 4 strains compared with 22.5 μg for 3 strains in the TIV). Furthermore, the safety profiles of the two vaccine groups were comparable. The results of this study demonstrated that the additional B strain in QIV did not compromise safety compared with TIV. These findings are similar to those of the meta-analysis, which did not find any significant differences in the systemic events, headache, and myalgia between the two vaccines [16].

In this study, immunogenicity was only assessed 28 days after the last vaccination. Hence, we do not know the duration of antibody responses to the 4 strains. Further study is needed to evaluate the antibody persistence of the QIV.

## Conclusion

In conclusion, this study demonstrated that QIV can be reproducibly manufactured to yield a well-tolerated, safe, and immunogenic vaccine in people aged 9–40 years and that it met all EMA immunogenicity criteria in adults. In adults, inactivated QIV induced comparable immune responses to TIV for A strains and the B lineage common to both QIV and TIV.

These data support the use of Bio Farma QIV for seasonal vaccination in children and adult subjects, which may enhance the protection against influenza and decrease the burden associated with influenza complications. The immunogenicity of the three batches of QIV was equivalent for all four strains.

## Data Availability

All relevant data are within the manuscript

## Acknowledgment

We would like to thank Dr. Iskandar, the Director of Bio Farma, for supporting this study. Thanks to Dr. Novilia S. Bachtiar, dr., M.Kes who has passed away for her contribution during various stages of the paper preparation. We would also like to thank the participants, Rita Verita Sri Hasniarty, the Head of Bandung District Health Office, Nitta Kurniati, the Head of Garuda Primary Health Center, and her staff, Siti Nurhasijatiningsih, the Head of Ibrahim Adjie Primary Health Center, and her staff, and Slyvie Virginati, the Head of Puter Primary Health Center, and her staff for their work in this study. We also thank Mr. Hadyana Sukandar for his statistical work in this study and acknowledge the Indonesian National AEFI Committee as the auditor of serious AEs in this study. We would also like to express our appreciation to the staff at the Clinical Research Unit of Growth Development-Social Pediatric Division for the invaluable administrative assistance.

## Contribution statement

**Eddy Fadlyana:** Conceptualization, Supervision, Data curation, Formal analysis, Funding acquisition, Investigation, Methodology, Project administration, Resources, Visualization, Writing – original draft, Writing – review & editing. **Meita Dhamayanti:** Conceptualization, Supervision, Data curation, Formal analysis, Investigation, Methodology, Resources, Writing – review & editing. **Rodman Tarigan, Susantina Prodjosoewojo, Andri Reza Rahmadi:** Conceptualization, Data curation, Formal analysis, Investigation, Resources, Writing – review & editing. **Rini Mulia Sari:** Conceptualization, Data curation, Funding acquisition, Methodology, Project administration, Resources, Validation. **Kusnandi Rusmil:** Conceptualization, Data curation, Formal analysis, Funding acquisition, Investigation, Methodology, Project administration, Resources, Supervision, Validation, writing – review & editing. **Cissy B. Kartasasmita:** Conceptualization, Methodology, Writing – review & editing.

## Ethics approval and consent to participate

This trial was approved by Research Ethics Committee Universitas Padjadjaran, Bandung (no.887/UN6.C.10/PN/2017). Written informed consent was obtained from the child’s parents and adult participants before performing any study procedure.

